# The role of fathers in feeding, care, and dental hygiene practices of children aged <6 years: A rapid scoping review

**DOI:** 10.1101/2024.03.19.24304543

**Authors:** Dina Moboshir, Priyanka Patil, Subarna Chakraborty, Joanna Dwardzweska, Clare H. Llewellyn, Kelley Webb-Martin, Carol Irish, Mfon Archibong, Jenny Gilmour, Phoebe Kalungi, Neha Batura, Monica Lakhanpaul, Michelle Heys, Logan Manikam, the NEON Steering Team

## Abstract

**Background:** The contribution of fathers in the early stages of child development, especially in feeding care and dental hygiene practices, is increasingly recognized but not well-documented. This rapid scoping review aims to broadly map the existing literature on this subject, focusing on children aged less than six years, and to identify areas where further research is needed.

**Objectives:** To explore the extent and nature of research on fathers’ roles in feeding care and dental hygiene practices for children under six years old. The review aims to identify key themes, variations in fatherly involvement across different contexts, and gaps in the current literature.

**Methods:** A structured search of key databases, including PubMed, PsycINFO, and Scopus, was performed. Studies included in the review involved fathers of children aged less than six years and addressed aspects of feeding care or dental hygiene. The process involved screening for relevance, categorizing studies into thematic areas, and summarizing overarching themes.

**Results:** The search yielded 15 studies encompassing diverse geographical and socio-cultural contexts. The review underscores the variability in fathers’ roles, influenced by factors such as urbanization, education, and cultural norms. It reveals that while fathers often serve as financial providers and role models, direct involvement in feeding and dental hygiene is less common. Notably, the literature on fathers’ involvement in children’s dental hygiene is limited.

**Conclusions:** Fathers’ roles in feeding care and dental hygiene practices for children under six years are multifaceted and context dependent. The review highlights significant gaps, particularly in understanding the involvement of fathers in dental hygiene. Addressing these gaps through future research is essential for developing comprehensive family-centred health care strategies and policies.

## Background

The parental influence on children’s feeding and dental health has been found to begin as early as during pregnancy [1], with high prenatal maternal intake of calcium being thought to improve foetal brain development through both indirect and direct means [2]. Prenatal maternal calcium intake has further been associated with risk of dental caries in the offspring via an inverse dose_response relationship between mothers’ prenatal intake of cheese and infant risk of dental caries [3]. Not only are mothers implicated in the feeding and dental hygiene habits of their infants, but fathers also play a major role, especially in determining whether a baby is breastfed exclusively [4]. The long-term impact of this can influence offspring health throughout the life course and even increase the risk of developing conditions such as cardiovascular disease [5], and yet while it has been suggested that shared parental involvement in such practices is more likely to result in healthier children, fathers are greatly underrepresented in this field of study and often even dubbed as the ‘invisible’ parent [6], as almost half of all studies regarding parenting and child health include only mothers as participants [7]. The reason for this rapid review focusing on feeding, care and dental hygiene practices as opposed to other aspects of parenting is due to the increased global prevalence of childhood obesity and dental caries. The World Health Organisation [8] reported that over 38 million children below the age of five years old were considered overweight or obese in 2019, and over 500 million children suffered from dental caries.

This topic has important implications for public health, as it is vital that healthcare officials look further into all possible reasons why there are inequalities in child health. This is particularly important in the case of conditions such as obesity and dental caries, which have an impact on children even after they have grown to become adults. Not only do obesity and dental caries have independent risks, but it has also been reported that there is a positive association between the two conditions, with a systematic review revealing that children with obesity had higher dental caries scores than those without obesity [9]. It is essential that public health officials investigate all causes and means in which parents can be supported to ensure that all children have equal starts to life, which includes investigating the role that fathers may play.

As society is becoming increasingly diverse regarding family structures, it is important for there to be further research conducted on the relations between father and child – this is clear as there have been studies that show the detrimental effects of father absenteeism on child development. It has been found that not only are behavioural issues at school more likely to occur in children who live without their fathers [10], but these children are also more likely to perform lower on academic tests than children who live with their fathers [11]. These findings make it clear that fathers must be encouraged to be involved in their children’s lives and that more awareness should be raised of the importance that fathers play in the lives of their children.

Studies that analyse the influence of parents on children’s development, eating habits and dental hygiene largely focus on the role of mothers and often neglect the importance of fathers within parenting. Studies also often focus only on childcare about eating habits and breastfeeding and do not consider the importance of parents in developing the basis for healthy lifelong dental hygiene habits. This rapid review aims to summarise the nature of studies on this topic and will be guided by the following research question:

⍰ “What is the role of fathers in the feeding, care, and dental hygiene practices for children under the age of six years?”

## Methods

### Search strategy

Rapid review methodologies were guided by the WHO’s practical guide to rapid reviews [43]. A systematic search was performed solely in the month of January 2023 through the following electronic databases: MEDLINE, Embase (1974–2023), APA PsycInfo and Global Health (1973–2023). All databases were searched through the Ovid platform. The search strategy involved looking for relevant studies that had the following keywords in their titles and/or abstracts: (father OR paternal) AND (infant or child*) AND (diet OR feeding OR eating OR nutrition) OR (dental OR oral hygiene OR oral health OR tooth brushing OR caries) OR (care). The results were limited to English language studies that were published between 2012–2022 and included only human participants. The combined number of studies retrieved totalled 831 (MEDLINE: 254, Embase: 351, PsycInfo: 109, Global Health: 117). All studies were downloaded into EndNote and screened for duplicates. The deduplication process resulted in the removal of 292 studies, amounting to a total of 539 studies. Once the titles and abstracts of each study were screened and compared against the inclusion and exclusion criteria, the total number of eligible studies was reduced to 45 studies. Full text copies of the eligible studies were obtained and screened using the same inclusion and exclusion criteria.

A supplementary hand-search was conducted on Google Scholar using the terms ‘father tooth brushing’ to account for the lack of eligible studies on the father’s role in dental hygiene practices that had reached the full text screening stage – this led to a total of 55 full texts being screened. After review of each full text, 32 studies were excluded, primarily for including participants beyond the age range specified or not specifically focusing on the aspects of fatherhood specified in the inclusion criteria (feeding, care, and dental hygiene). This resulted in 23 eligible studies (11 mainly discussing feeding, 7 mainly discussing care, and 5 mainly discussing dental hygiene). Five most recently published relevant studies about each of the subtopics (feeding, care, and dental hygiene) were selected to be a part of the final sample of studies. These studies were chosen per subtopic so that our review could accurately understand the scope of studies in these subtopics regardless of whether the field of research on a particular subtopic was scarce. We chose to specifically include the most recently published articles to accurately reflect the rapidly changing nature of societal family dynamics. Additionally, the five most recent publications chosen led to ensuring a through, critically appraised quality and relevance of each study, aided in balancing perspectives to account for differing conclusions and methodologies, and ensure better comparability across the subtopics. A rapid review methodology was also chosen due to the restricted time constraints of the dissertation project. Supplemental Material 1 presents a PRISMA (Preferred Reporting Items for Systematic Reviews and Meta-Analyses) flow diagram that outlines the study selection process.

### Inclusion and exclusion criteria

The review incorporated studies that met the following inclusion criteria: English language publications from 2012-2022, involving fathers of children aged 0-6 years at the study’s time, with a distinct emphasis on child feeding, dental hygiene practices, and/or childcare. This age range was selected to specifically scrutinize research on fatherhood during infancy and early childhood. To comprehensively analyse the research scope, methodologies encompassed quantitative, qualitative, and mixed methods designs.

Exclusion criteria comprised studies published before 2012, those focusing on single-father households, non-human studies, and those without publicly available full texts. Single fathers were deliberately excluded due to the unique family dynamic introducing additional confounders and presenting an inequitable comparison with dual-parent households.

### Data extraction

Relevant data were retrieved from the full texts of all 15 studies and compiled into an Excel spreadsheet. The following data were extracted from each study: citation, article type, aims and objectives, study design, location and date of the study, participant demographics, recruitment techniques, sample size, details of intervention (if applicable), data collection method, losses to follow-up, key results, and limitations. An overview of this is provided in Supplemental Material 2, along with the quality assessment scores.

### Quality assessment

To streamline the review process according to the WHO practical guide to rapid reviews [43], a single reviewer (DM) conducted quality assessment of the studies. The Critical Appraisal Skills Programme (CASP) ten-question qualitative studies checklist [12] was used to assess the methodological quality of qualitative studies (of which there were seven [14, 16–19, 24, 26]), and the 12-question cohort study edition was used for one study [20]. The Strengthening the Reporting of Observational Studies in Epidemiology (STROBE) 22-item cross-sectional study checklist [13] was used for all studies with a quantitative component, of which there were seven [15, 21–23, 25–28]. The studies were scored with a system of the answer ‘Yes’ accounting for one point; therefore, qualitative studies had a maximum score of 10, quantitative studies had a maximum of 22 and the single cohort study had a maximum of 12. No studies were excluded for being of inadequate quality.

## Results

The final sample included 15 studies that were conducted in nine countries, predominantly in Asia (India, *n*=2; Pakistan, *n*=1; Iran, *n*=1; China, *n*=1) and North America (US, *n*=5; Canada, *n*=1), with the remaining (*n*=4) being from Europe (*n*=1), Africa (*n*=2) and one global online survey. Studies were conducted in both urban and rural settings. In total, 7,689 parents were studied, with sample sizes ranging from 18 [17] to 2,229 [28] participants. Participants were recruited from a variety of settings, including hospitals (*n*=6), day-care centres (*n*=4), and online (*n*=2). Several different data collection methods were used, including interviews [16–20, 22, 26], focus group discussions [14, 19], questionnaires [15, 16, 20–23, 25–28], observations [24, 26], physical assessments [25] and cognitive assessments [26].

Based on the fifteen studies reviewed, six common themes were identified: fathers as providers of food, seeking healthcare as a shared parental role, fathers as role models, father’s role in play, fathers as supervisors and supporters of mothers as well as urban and rural differences.

### Fathers as providers of food

The view of fathers as providers of food was cited in multiple studies [14, 18, 19]. Both mothers and fathers reported that although fathers provided the financial provision to buy meals, mothers are expected to prepare and feed the food to their children [14]. Fathers often reported believing that mothers were more aware of the child’s specific needs, likes, and dislikes due to mothers spending more time at home [19]. Awareness of obesity was reported as a common factor that influenced fathers’ involvement in the feeding of their child, as fathers reported wanting to encourage their child to consume more fruit and vegetables to reduce the risk of the child developing childhood obesity [18]. Jeong et al. also found that many fathers prioritised purchasing fruit, vegetables, and milk for their children to grow healthy [19]. However, this view was not consistent across all studies, as Schoeppe and Trost reported that paternal support, as measured by behaviours such as preparing fruits for their child and consuming fruits and vegetables together with their child, was not associated with children’s total fruit and vegetable intake [25]. One study found that fathers reported feeling guilty after giving in to their child’s requests for unhealthy snacks despite being aware of the potential health consequences associated with poor nutrition [18]. The dietary diversity of children was significantly associated with several different predictors of fathers’ knowledge, including knowledge of food groups, childcare and health-related knowledge [16]. Children with high rates of dietary diversity were more likely to have fathers who were considerably involved in daily childcare activities [16]. Breastfeeding was reported to be a barrier for fathers who wished to feel more involved in infant feeding and care [15, 19]. Fathers of infants generally reported believing that breastfeeding was the best option for their child [15, 19], but many felt that it reduced their role as a parent [15]. The introduction of bottle-feeding alleviated some of these feelings, as fathers reported feeling more able to bond with their child and divide childcare more equally between parents [15]. Several mothers reported that fathers ordered them to feed their child in ways that were not in line with recommendations from their healthcare providers [14].

### Seeking healthcare as a shared parental role

The concept of children’s health being a shared responsibility between mother and father was reported in several studies [18, 19]. Parents described attending healthcare visits, purchasing medicine, and administering medication together for their child [19]. Gender patterns were identified within this shared role, as parents reported fathers primarily being responsible for physically taking the child to healthcare visits as well as providing the financial means to attend the visits and purchase medicine [19]. This contrasts with the mother’s described responsibility of administering medication and observing the child’s symptoms and progress [19]. Garfield and Isacco reported that two-thirds of fathers were comfortable in administering medication to their child, but several fathers were not confident in administering the correct dosage or preferred giving as little medicine as possible to their child [18]. Several fathers reported feeling doubtful in their role as a primary caregiver when the child’s mother was not present during emergency care visits [18]. Mothers were shown to have a significantly greater mean knowledge of infant dental care than fathers [21], with it being found that mothers were the most likely to first introduce toothbrushing to their child [22]. This finding was also reported by another study that found that fathers had a much lower level of child oral health knowledge than mothers despite most fathers stating that they had an average level of knowledge [27]. Pullishery et al. found that only mothers’ and not fathers’ preventive dental care influenced children’s oral health [22]. In contrast, Shaghaghian and Zeraatkar reported a univariate analysis that found a significant association between child frequency of toothbrushing, fathers’ level of education and fathers’ employment [23]. The same study reported that fathers’ education was significantly associated with children beginning toothbrushing before the age of two years old; however, these effects were not consistent in multivariate analysis [23].

### Fathers as role models

Multiple studies implicitly portrayed fathers as role models for their young children [19, 24, 26, 28]. A study based in Pakistan spoke of how fathers bonded with and taught shared values to their children through religion by taking them to the mosque and encouraging them to learn how to recite the Qur’an [19]. Participants were very aware of how children model their own behaviour based on the behaviours exhibited by both of their parents [19]. Fathers reported wanting to display healthy behaviour for their child to learn from, such as eating vegetables and drinking milk at the dinner table and reducing the amount of alcohol they drink [18]. Promoting good hygiene practices was seen as another way in which fathers demonstrate healthy behaviours for their children to model [18]. Saltzman et al. reported that children who had fathers present during dinner time were less likely to be distracted by analogue objects such as books and toys than children whose fathers were unavailable during dinner time [24]. Fathers’ presence during dinner time was also found to influence mothers’ behaviour, as mothers were more likely to engage in responsive feeding practices, such as providing positive food-related interactions and giving their child food options to choose from if the father was present [24]. Feeley et al. found that the fathers who were the most involved in direct caregiving of their child were those who believe that optimal child development requires the involvement and guidance of fathers as well as mothers [17]. This view is supported by another study that found that higher levels of emotionally supportive paternal behaviour were associated with children’s greater executive functioning at 24 months old [26]. This association was not present at 7 months old, suggesting that the father’s role may become more prominent during toddlerhood [26]. A study with a large sample size of 2,086 parents found that families in which the father was the main caregiver were less likely to report experiencing child-rearing difficulties such as feeding, sleeping and developmental problems [28].

### Father’s role in play

Many studies have emphasised how fathers felt as if playing with their children was one of their most important roles as a father [18, 19, 20]. Garfield and Isacco found that almost half of all fathers in their study reported exercising as a means of modelling and promoting healthy behaviour for their children [18]. Fathers described exercising with their child through activities such as ball games and cycling as a way of bonding and strengthening their father-child relationship [18]. This was a consistent theme across several studies, as Jeong et al. reported that fathers expressed playing games such as hide-and-seek and marbles with their child as not only a method of promoting their child’s physical and cognitive development but also as a means of expressing love and affection [19]. A child gender division was reported, as fathers of sons were found to spend more time engaging in physical play with their child compared to fathers of daughters, who were more likely to participate in literacy activities instead [20]. Many parents reported that fathers also often occupied the role of taking their children outside for outings, such as visiting relatives and exploring [19]. Parents explained how children asking questions because of their natural curiosity about their surroundings enabled fathers to help children develop their knowledge and memory as well as allowing for children to experience new environments by leaving their homes [19]. Schoeppe and Trost found that paternal (and maternal) support for children’s physical activity, as defined by factors such as playing outside together with your child and supporting your child in participating in sports, was positively associated with the physical activity levels of preschool-aged children [25]. Fathers’ roles as providers are also present in the context of play and physical activity, as a study reported that fathers were expected to pay for all aspects of childcare, including play materials [19]. Despite this cost, fathers reported enjoying seeing their child happy playing and exploring during outings and considered this to be a mutual feeling, as their children were often the ones to initiate the outings and request for their fathers to take them outside [19].

### Fathers as supervisors and supporters of mothers

Several fathers reported feeling that their role as a parent included supporting and encouraging the mother of the child so that the child would grow up in a healthy and nurturing environment [14, 15, 17]. This was particularly the case when infants were being breastfed, as many fathers felt that their interactions with their child were limited, so it was important for mothers to be supported to develop a strong mother-child bond [15, 18]. Several fathers described how they believed that, during infancy, the mother-child bond is more important than the father-child bond and that fathers should work as part of a ‘chain’ and take care of other elements of family life so that the mother could tend to the infants’ needs [17]. These fathers described working, tending to household chores, and taking care of older siblings as factors that they undertook to support the mother [17]. This view of fathers doing household chores was not consistent across all studies, as other studies reported fathers viewing household chores as mothers’ responsibility due to them spending more time at home; however, clear distinctions between mothers’ home responsibilities and fathers outside responsibilities were made [18]. The influence of fathers on mothers’ behaviour was reported by Saltzman et al., as a bivariate analysis revealed that mothers were more likely to spend a smaller proportion of time engaged in distractions if fathers were present during mealtimes [24]. Some fathers were much less enthusiastic about the reverse role of mothers as supporters of fathers and expressed clear displeasure, with one participant referring to the role as if ‘*the sky is now on the ground’* [14]. This displeasure was combined with the description of fathers playing a supervisory role by monitoring mothers in the home to ensure they were fulfilling their role as a primary caregiver [14]. Fathers who viewed childcare and parenthood as a more egalitarian shared role between mother and father had an increased likelihood of high paternal involvement [15].

### Urban and rural differences

As the studies consisted of participants from both urban and rural settings, a clear difference in views became evident. Many fathers in rural settings spent a limited amount of time at home, as the most common occupations in these settings involved agriculture and physical labour jobs with long working hours, often in areas that were far from the fathers’ homes [19]. This led to many fathers migrating away from their families to seek higher-paying jobs [19]. A consistent theme among studies that reported data from both urban and rural settings was that families from rural settings often expressed more traditional views towards gender roles and parenthood [14, 19]. Rural families were more likely to describe having distinctly separate roles as parents, with fathers being the sole breadwinners and mothers being the primary caregivers of children [14]. Fathers from rural areas often reported feeling as if they were lacking as a parent due to financial struggles not enabling them to easily attain all their child’s needs and requests and fulfil their role as a provider and breadwinner [19]. The importance of traditional values in rural settings is evident, as these gender roles were persistent even when extended family members were helping with childcare, as aunts and grandmothers were reported to support mothers in their household and childcare duties, while uncles supported fathers in playing with the child and taking them to outings [19]. In comparison, mothers and fathers from urban settings reported less traditional and more liberal approaches to parenting. Several mothers from rural settings described how rural fathers were less helpful in childcare and parenting than those from urban settings [14]. Urban fathers reported more instances of decision-making with mothers to equally divide parenting roles and were more likely to engage in completing household chores and buying nutritious food for not only their children but also their mothers [14]. Fathers’ level of education was also important in influencing how they interact with their children, as more educated fathers reported the importance of a balanced diet and nurturing environment for children’s early development [19]. Education was important in determining how parents divided parenting roles, as several mothers from rural areas reported having to stay at home and oversee household chores and childcare because they were illiterate [19]. Higher parental levels of education and socioeconomic status were also consistently reported to be associated with an increased level of child oral health knowledge [21–23].

## Discussion

The findings of this rapid scoping review contribute to the growing body of evidence highlighting the importance of fathers in the feeding, care, and dental hygiene practices of their children. Six themes were identified from the fifteen academic papers that were studied: fathers as providers of food, seeking healthcare as a shared parental role, fathers as role models, fathers’ role in play, fathers as supervisors and supporters of mothers, and urban and rural differences in parental roles. Overall, fathers play a larger role in children’s early development than once thought, but these influences differ in urban and rural settings.

### Feeding

Most studies that investigated the father’s role in feeding found that the most common route in which fathers influenced their children’s feeding was through providing the financial means of purchasing food [14, 18, 19]. In both urban and rural settings, the action of feeding the child was often portrayed as the mother’s duty; however, urban fathers had more instances of reporting wanting to be involved in feeding. This view is consistent with previous research that has found a common theme of the role of feeding being seen as a maternal role with fathers helping only if necessary [29, 30]. Rather than feeding the child or preparing meals themselves, a pattern of fathers reporting being involved in promoting healthy foods through modelling behaviour was evident. One father described this as *‘monkey see, monkey do,’* and this need to promote healthy eating was particularly important for fathers in the context of rising obesity epidemics in their country [18]. Previous research has shown that fathers may be more likely to feel guilty and give in to unhealthy food requests from their child [31, 32], and this is in line with studies analysed in this review that reported fathers buying unhealthy snacks for their children [18, 19] and often feeling guilty afterwards [18]. This review also unveiled how, despite supporting breastfeeding, many fathers felt as if they were the secondary parent when their child was reliant on breastfeeding [15, 19], and this is consistent with other studies that have reported fathers seeing breastfeeding as a barrier to them forming a bond with their infant [33]. This suggests that the father-child relationship is particularly important during toddlerhood once children cease being breastfed and aligns with studies in this review that reported fathers feeling that their bond improved once children began being bottle-fed and eating solid foods [15].

### Care

This review revealed how many fathers see play and outside time as their greatest contributions to childcare. This was consistent across both urban and rural settings, as fathers stressed how playing and outings were not only enjoyable for both parents and children but were also important in helping the child ‘freshen’ their mind and learn more about the outside world [18, 19, 20]. This importance was clear, as fathers reported that male relatives often took over this role if the father had migrated away for work or was preoccupied [19]. This is in line with prior research, as Kazura found that children presented higher play scores and played at higher levels in father-child dyads than in mother-child dyads [34]. This review also found that fathers’ level of involvement often depended on the age and gender of their child [19, 20]. Leavell et al. reported that this gender difference became particularly evident after children reached the age of two years old as fathers of sons began to engage more in physical playing compared to fathers of daughters engaging more in reading and telling stories [20]. Several families of multiple children also mentioned how fathers were more comfortable undertaking caregiving activities such as bathing their child with sons rather than daughters [19]. This supports previous research that found that across all age groups, both mother and father were more likely to bathe with children of the same gender as them [35]. Fathers have also been found to be less likely than mothers to participate in physical care tasks generally, with half of mothers’ childcare time being allocated to this compared to a third of fathers’ childcare time [36]. Overall, the level and type of interactions that fathers had with their children greatly reflected the wider society and urban/rural environment context in which the studies were conducted. The influence of gender roles and fathers’ occupations were very apparent across all studies, as the behaviours of fathers seemed to be highly reliant on what they viewed as mothers’ and fathers’ duties.

### Dental hygiene practices

The study search process of this review highlighted the lack of research surrounding the influence of fathers on their children’s oral health and toothbrushing habits. Across all studies within this review that investigated parents’ role in children’s oral health, fathers showed a clear lack of oral health knowledge compared to mothers [21, 22, 27]. This supports previous research that also found that fathers had a lower level of dental care knowledge than mothers [37]. Despite this, fathers’ level of education has been found to influence children’s oral hygiene habits both within this review [23] and in previous studies in which children of fathers with high educational levels were more likely to have a greater frequency of toothbrushing [38]. It has also been reported that fathers’ greater frequency of toothbrushing is associated with children’s lower scores on the Decayed, Missing due to caries, and Filled Teeth (DMFT) index [39]. However, this finding has not been consistent across previous studies, as Karaaslan et al. found that while parental educational level was associated with children’s frequency of dentist visits, there was no significant association between parental educational level and child toothbrushing frequency [40]. This suggests that further research must be done on this topic to clarify the relationship between parental education and children’s oral health and toothbrushing habits.

### Strengths and limitations

This rapid scoping review aimed to provide an overview of the ways in which fathers interact with their children through feeding, care, and dental hygiene. A strength of this review was that the inclusion of studies based in both urban and rural settings and countries across the globe allowed for the recognition of how context and societal norms greatly influence the roles fathers play in their children’s lives. The mixed methods nature of the review also allowed for the analysis of both quantitative and qualitative studies, which enabled a better depth of understanding of the role fathers play. This systematic review had several limitations. First, as there was a lack of available resources on fathers’ impact on child oral health, a manual search of Google Scholar was necessary, which introduced the risk of selection bias. This lack of resources also limits the extent to which the findings can be to other topics related to father-child interactions. Second, the review was limited to the fifteen most recent studies available; therefore, many relevant studies may have been excluded, so the review may not accurately reflect the complexity and diversity of father-child interactions. Third, the variable methodological quality of the studies reduces the reliability and validity of the findings, and the use of a more rigorous quality appraisal process would have been beneficial. Finally, the wide usage of self-report measures may cause the data to be prone to bias and limit the accuracy of the data. The nature of the topic of the role’s fathers play in the lives of children makes the studies vulnerable to social desirability bias, which may influence the validity of the findings.

### Implications

These findings support the growing body of evidence highlighting the importance of the role fathers can play in their children’s lives. Healthcare professionals and teachers should work to involve both mothers and fathers in conversations involving their children. The involvement of fathers in supporting breastfeeding should be further encouraged by nurses and prenatal support groups, as this study supports the view that fathers often feel discouraged as parents and unable to bond with their children during this infancy period. This is particularly important to address, as research has found that paternal support of breastfeeding has a strong impact on the likelihood and duration of mothers breastfeeding [41]. Interventions to increase breastfeeding rates that target both mothers and fathers have also been found to be more effective than those that target mothers alone [42]. Further research should be conducted around fatherhood overall, but this review also highlights the importance of investigating the role of fathers in child oral health in further depth, as there is mixed evidence on this topic.

## Conclusion

The purpose of this review was to gain further insight into fatherhood, and recent literature supports the view that fathers play various important roles in the lives of their children, with strong influences on their children’s feeding, care, and dental hygiene habits. The review has highlighted how fathers can influence children by being providers, playmates, and role models as well as through healthcare. The review emphasises the importance of coparenting and sheds light on the positive and negative influences that fathers can have on mothers. A number of gaps in research were found in the process of conducting this review, and several issues in need of further investigation were evident: a) few studies investigated fathers’ influence on dental hygiene habits in depth, and there is inconsistent evidence on the exact impact fathers have, b) several papers unveiled fathers’ differential gender treatment of their children, and c) there are clear urban and rural differences in fatherhood. Further support should be given to encourage fathers to be more involved in the lives of their children, particularly during infancy – this could be in the form of providing fathers with informational leaflets or tailored parenting classes specifically targeting fathers. The review highlighted a gap in fathers’ knowledge of child oral health, and there should be both further research on this overall topic and an increased emphasis on encouraging fathers to learn more about the importance of toothbrushing, dietary habits and regular dentist visits. The findings of the review have implications for public health, as they suggest that public health researchers and policymakers should consider ways to make father involvement seem more appealing through policies such as paid paternity leave.

## Data Availability

All data produced in the present study are available upon reasonable request to the authors

## Acknowledgements

The authors would like to acknowledge the contribution of the NEON steering team in the development of this scoping review. Members of the NEON steering team consist of Dr Logan Manikam, Dr Priyanka Patil, Prof Michelle Heys, Prof Monica Lakhanpaul, Dr Neha Batura, Prof Atul Singhal, Prof Mitch Blair, Kelley Webb Martin, Carol Irish, Dr Mfon Archibong, Joanna Drazdzewska, Dr Sonia Ahmed, Amelie Gonguet, Gary Wooten, Dr Ian Warwick, Vaikuntanath Kakarla, Phoebe Kalungi, Jenny Gilmore, Prof Richard Watt, Prof Audrey Prost, Dr Edward Fottrell, Ashlee Teakle, Prof Oyinlola Oyebode, Dr Keri McCrickerd, Dr Rana Conway, Professor Lisa Dikomitis, Mari Toomse-Smith, Scott Elliot, Julia Thomas, Aeilish Geldenhuys, Chris Gedge, Kristin Bash, Dr Dianna Smith, Kate Questa, Dr Megan Blake, Gary Tse, Dr Queenie LAW Pui Sze, Gavin Talbot, Dr Chiong Yee Keow, Dr Angela Trude, Prof Lindsay Forbes, Dr Nazanin Zand, Dr Clare Llewellyn, Lakmini Shah, Subarna Chakraborty, Yeqing Zhang, Sumire Fujita, Dina Moboshir, Natasha Chug, Tala Khatib and Delaney Douglas-Hiley.

Steering team members had an opportunity to critically review results and contribute to the process of finalising this paper.

The authors would like to thank the National Institute of Health Research, Collaboration for Leadership in Applied Health Research and Care North Thames for funding the NEON study. This work is supported by the NIHR GOSH BRC. The views expressed are those of the author(s) and not necessarily those of the NHS, the NIHR or the Department of Health.

## Author’s Contributions

DM, PP and SC contributed to the manuscript writing, and prepared it for submission. DM, PP, ML and LM had primary responsibility for the final content. All authors read and contributed to reviewing the study data, the designing of the manuscript, and the approval of the final manuscript.

## Disclaimer

The views expressed in the publication are those of the author(s) and not necessarily those of the sponsor (UCL), funder (NIHR), study partners (Tower Hamlets GP Care Group, London Borough of Newham Council).

## Participant consent for publication

Participant information sheets and consent forms were provided to community members and those expressing interest. Community participants agreed to participate and gave audio/video consent prior to their participation in the workshops. Verbal consent was witnessed and formally recorded. All participants were informed of their right to freely withdraw from the study at any time. Confidentiality of personal data was ensured using anonymisation techniques as stated in the Data Protection Act (1998) and in line with the General Data Protection Regulation (2018). All participant data is anonymised and stored on an encrypted password protected computer. Data can only be accessed by the authorised research personnel.

## Ethics approval

This study has obtained ethical approval from UCL Research Ethics Committee [Ethics ID 17269/001], Sponsor reference number: 142600, Funding Reference: NIHR300020 and IRAS number: 296259, Ref: 21/SW/0142. Study protocols and relevant documents were reviewed by UCL Research Ethics Committee, NHS Health Research Authority (HRA) and study partners involved in establishing data sharing agreement for linking participants’ routine data (Tower Hamlets GP Care Group, London Borough of Newham Council).

## Provenance and Peer Review

Not commissioned; peer reviewed for ethical and funding approval prior to submission.

## Data Sharing Statement

The data supporting the findings of this study is available upon reasonable request from the corresponding author.

## List of abbreviations

*PRISMA*: Preferred Reporting Items for Systematic Reviews and Meta-Analyses
*DMFT*: Decayed, Missing due to caries, and Filled Teeth
*CASP*: Critical Appraisal Skills Programme
*STROBE*: Strengthening the Reporting of Observational Studies in Epidemiology
*US*: United States of America

## Supplemental Materials

**Supplemental Material 1:**
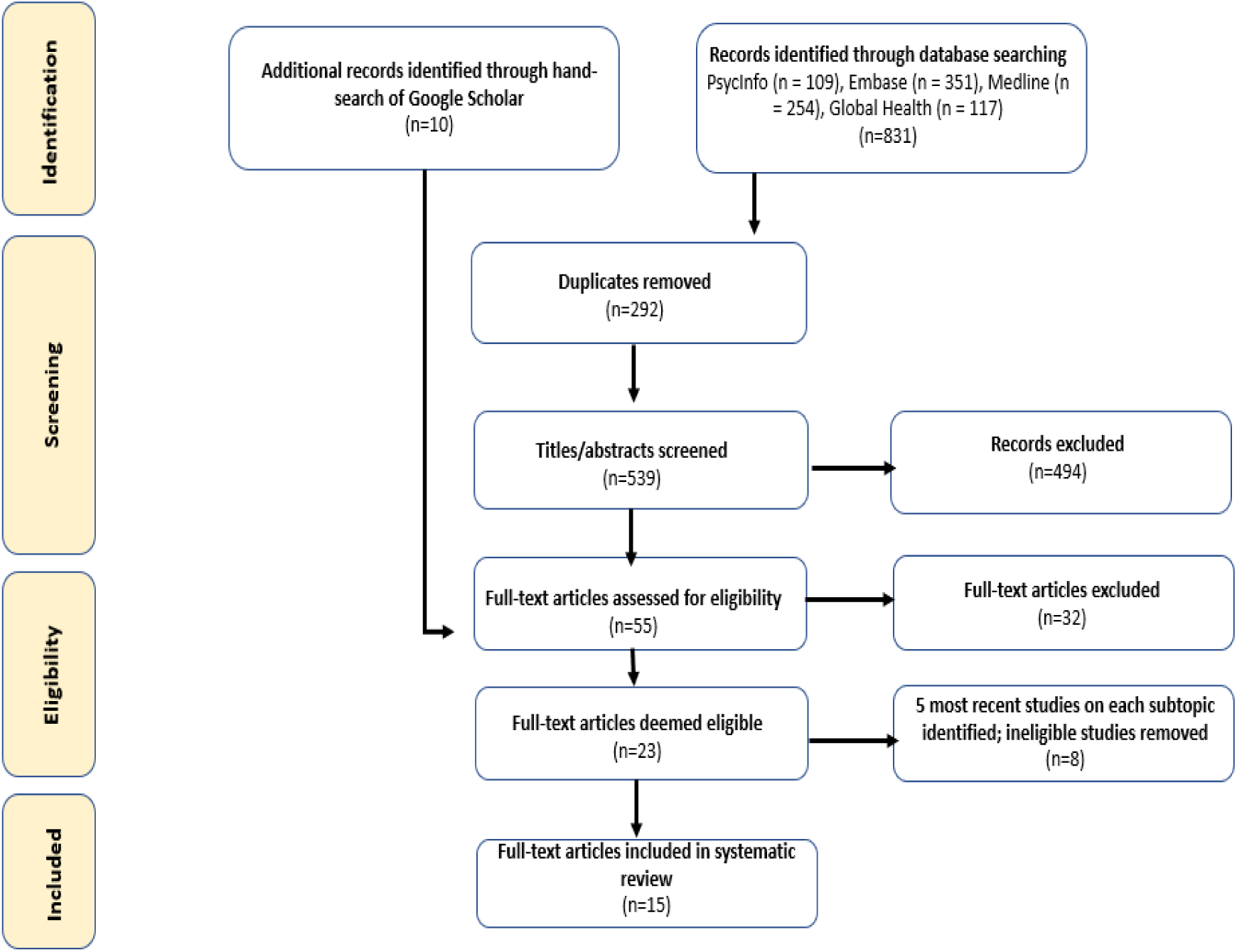
PRISMA flow diagram of the search strategy results.

**Supplemental Material 2:** Summary of data extraction and quality assessment.

| Reference | Author,<br>publication<br>year | Location | Sample size | Methods | Quality<br>assessment<br>score |
| --- | --- | --- | --- | --- | --- |
| 14 | Allotey et al. (2022) | Nigeria | 160 total mothers and fathers | Focus group discussions (FGD) | 8/10 |
| 15 | Atkinson, Silverio and Fallon (2021) | Online | 212 participants | Online survey | 16/22 |
| 16 | Bilal et al. (2014) | Ethiopia | 850 mothers, 850 fathers | Questionnaire conducted by interviews | 9/10 |
| 17 | Feeley, Sherrard and Boisvert (2013) | Canada | 18 fathers, 21 infants | Interviews | 9/10 |
| 18 | Garfield and Isacco (2012) | United States of America (US) | 31 fathers | Interviews | 10/10 |
| 19 | Jeong et al. (2018) | Pakistan | 40 fathers, 39 mothers | Interviews and FGD | 10/10 |
| 20 | Leavell et al. (2011) | US | 426 fathers, 426 children | Questionnaire conducted by interviews | 10/12 |
| 21 | Nagarajappa et al. (2013) | India | 470 participants | Self-administered questionnaire | 14/22 |
| 22 | Pullishery, Shenoy and Shenoy (2013) | India | 127 caregivers, 130 children | Questionnaire conducted by interview | 14/22 |
| 23 | Shaghagian and Zeraatkar | Iran | 396 parents, 453 children | Self-administered | 16/22 |
|  | (2017) |  |  | questionnaire |  |
| 24 | Saltzman et al.<br>(2019) | US | 109 families | Observational<br>home visits | 9/10 |
| 25 | Schoeppe and<br>Trost (2015) | US | 346 parents,<br>173 children | Physical<br>assessment of<br>children and<br>parent self-administered<br>questionnaire | 19/22 |
| 26 | Towe-Goodman et<br>al. (2014) | US | 1,240<br>parents, 620<br>children | Observations,<br>interviews and<br>cognitive<br>assessments of<br>children | 7/10 |
| 27 | Wapniarska,<br>Bula and Hilt<br>(2016) | Poland | 146<br>participants | Self-administered<br>questionnaire | 12/22 |
| 28 | Zhang et al.<br>(2019) | China | 2,229<br>parents | Self-administered<br>questionnaire | 19/22 |

